# The Time Course of Clinical Oxygenator Failure Due to Clot Formation

**DOI:** 10.1101/2020.11.20.20235606

**Authors:** Caitlin T. Demarest, Samantha J. Shoemaker, Michael P. Salna, Scott R. Chicotka, Kenmond Fung, Matthew D. Bacchetta, James F. Antaki, Keith E. Cook

**Author notes:** Address for correspondence: Keith E. Cook, Ph.D., 4N207 Scott Hall, 5000 Forbes Ave, Pittsburgh, PA 15213.

## Abstract

**Background:** Long-term use of extracorporeal membrane oxygenation (ECMO) remains limited because of poor biocompatibility, which often leads to clot formation and device failure. Despite this common pathway to failure, there are no published studies on the rate of clot formation and resulting performance deficits in current oxygenators.

**Methods:** ECMO cases with either Maquet’s CardioHelp (CH, n=28) or Quadrox (Qx, n=14) oxygenators were evaluated over a three-month period. Data was collected prospectively and included patient characteristics and hematological data. The inlet-outlet oxygen content difference (ΔC_O2_) and blood flow resistance were calculated as measures of device function, and device failure due to clot formation was defined as a resistance increase greater than 1 mmHg/(L/min)/day for more than one day.

**Results:** There were no statistically significant differences in patient age, total days on ECMO, platelet count (PLT) prior to ECMO, activated partial thromboplastin time during ECMO, initial resistance, and device blood flow rate. During ECMO, the Qx group had a significantly greater change in PLT (Qx: - 34±10%; CH: 7±15%), rapidity to failure due to clot formation, and a greater decline in ΔC_O2_ (all p<0.05). Clot burden was focused at the center of the CH oxygenator, farthest from all inlets, whereas Qx devices developed a more diffuse clot pattern.

**Conclusions:** Qx oxygenators clot earlier than CH oxygenators with a correlated drop in ΔC_O2_ and greater PLT reduction. These differences are likely due to the distributed, four-inlet CH design vs. the single inlet Qx design and differences in pump-induced platelet activation.

## Introduction

Extracorporeal membrane oxygenation (ECMO) is being used with increasing frequency and duration for both respiratory and cardiac support as a bridge to recovery, durable device support, or transplantation (BTT).^1^ Despite increased usage and protocolized anticoagulation, ECMO oxygenators remain plagued by progressive performance deterioration and biocompatibility issues, including bleeding, clotting and thromboembolic complications.^2,3^ These limitations limit their effectiveness for long term BTT support and preclude their use as durable support devices for patients who are ineligible for lung transplantation. These patients continue to live with chronic lung disease, burdened by continuous home oxygen requirements, frequent and costly hospitalizations, and no restorative therapies.

The main factor preventing the long-term use of these devices in chronic lung disease patients is clot formation. As clot propagates, oxygenator gas exchange declines, blood flow resistance increases progressively and an injurious cycle of hemolysis can occur. Clotting is initiated at the artificial surface with protein adsorption, which then causes platelet adhesion and activation, initiation of the coagulation cascade, and subsequent thrombus formation.^4,5^ Although there is a breadth of research focused on improving anticoagulation and antithrombotic coatings, there are limited publications that track the time course of clot formation and device functional deterioration within the most commonly used clinical oxygenators. A few investigators have evaluated D-dimer, computed tomography, and pressure drop to evaluate clot formation and determine if a device is failing,^6-8^ but none has sought to connect these strategies to average device performance. Broader understanding of the location and time course of clot formation within these devices will allow engineers to analyze failure mechanisms and improve the design of future devices, while also giving clinicians important feedback into their current ECMO decision-making.

To analyze patterns of clot formation and device functional deterioration, clinical information was collected from all ECMO cases over a three-month period at Columbia University Medical Center. Every oxygenator was examined for physical and functional patterns indicating clot formation, whether removal was for device failure or patient weaning. The time course of functional deterioration in each device was determined using longitudinal changes in blood flow resistance and oxygen transfer. The probability of clot formation at each location in the gas exchanger inlet and outlet was also determined as a function of time using digital image processing, and the extent of clot was compared to the observed functional deterioration. Patient demographics and laboratory values were compared to assess their effects on the rate of deterioration. Results suggest significant differences in long-term performance between the Qx and CH systems.

## Methods

All ECMO cases were evaluated over a three-month period, regardless of the indication, length of ECMO course, or reason for discontinuation (i.e. weaning due to patient recovery, patient transplant, or device failure). All patients received a bolus of 5,000u heparin at the time of ECMO cannulation, and then were maintained on a heparin drip thereafter with a targeted and activated partial thromboplastin time (aPTT) of 40-60s. Patients were given packed red blood cells when their hemoglobin fell below 7, and a platelet count less than 50 was tolerated in absence of bleeding.

The ECMO systems at Columbia are either Maquet’s CardioHelp (CH) or a Quadrox (Qx) oxygenator coupled with either a Rotoflow or CentriMag pump. These two oxygenators both consist of polymethylpentene hollow fibers with Bioline heparin coating and a 1.8m^2^ gas exchange surface area. However, CH is an integrated pump-oxygenator with four inlets, whereas the Qx is a stand-alone oxygenator with one inlet that is paired with a pump. These subtle variances lead to different flow patterns within the devices, which might produce different patterns of clot formation. The type of device used on a patient was at the clinician’s discretion. However, a Qx was typically used if the patient was immobile and likely to remain so whereas a CH was used if the patient was expected to be ambulatory or be transported from an outside hospital to Columbia; this decision was based on the less expensive cost of the Qx versus the smaller size and fewer independent components of the CH system.

Patient demographics collected included age, indication for ECMO, and type of cannulation. Clinical parameters measured included arterial blood gases (ABG), hemoglobin (Hgb), platelet count (PLT), and aPTT. Circuit data was collected daily throughout the ECMO course, including device flow (Q), inlet and outlet pressures, inlet and outlet blood gases, and sweep gas flow rate. Statistical analyses were performed using SPSS software (SPSS, Chicago, IL). All data are presented as mean and standard deviation. The statistical significance of most variables was determined by the two-sample t-test assuming unequal variances. A p-value of < 0.05 was required for statistical significance. When comparing PLT counts between groups, patients that received platelet transfusions were excluded from platelet trends (CH n=5, Qx n=5) to remove it as a confounding factor. Device failure was defined as an increase in blood flow resistance greater than 1 mmHg/(L/min)/day for more than one day; survival analysis using this failure criterion was performed using the Kaplan Meier method. As a measure of device performance, the inlet-outlet oxygen content difference (ΔC_O2_) was determined (O_2_ content = hemoglobin x saturation x 1.34 x PaO_2_ x 0.003).

Following detachment from the ECMO circuit, the oxygenators were rinsed with saline until the effluent was clear and photographed. A program was developed to process these photographs and generate a probability density map of clot formation. To do this, the images were converted to grayscale, and a darkness threshold value between 129 and 255 was used to identify clot. The images were converted to binary using ImageJ, where a clot is depicted as white, and the remaining image is black. These binary images were then aligned and imported as a stack into MATLAB. The program evaluated each individual pixel in each image to generate a matrix of 0s and 1s. If the pixel is black, then the equivalent position in the matrix is left as 0, and if the pixel is white, then the equivalent position is assigned a 1. The program then sums each matrix and averages them, resulting in a scale from 0 to 1 at each pixel. The resulting image depicts the probability of clot formation on an absolute color scale, where 0 is blue (0% clot frequency), and 1 is red (100% clot frequency).

## Results

A total of 28 CH and 14 Qx cases were evaluated. **Table 1** summarizes selected pertinent data. There were no statistically significant differences in patient age or total number of days on ECMO. The CH patients were most commonly in pure respiratory failure needing VV cannulation, whereas the Qx group had a higher percentage of patients needing cardiopulmonary support with VA cannulation. There were no statistically significant differences between groups in mean platelet count prior to ECMO initiation or mean aPTT throughout the ECMO course, indicating similar coagulation profiles between the two patient populations. Additionally, there was no statistically significant difference in initial device resistance between the two groups, and there was no difference in average device blood flow rate throughout the ECMO course, indicating a similar amount of support delivered in each group.

Although there were no differences in their initial platelet count, the CH group had an average percent change in platelet count of 7.19 ± 15.45%, whereas the Qx group had a percent change of -33.87 ± 10.26% (p<0.05) by the end of the ECMO run (**Figure 1**).

**Figure 1:**
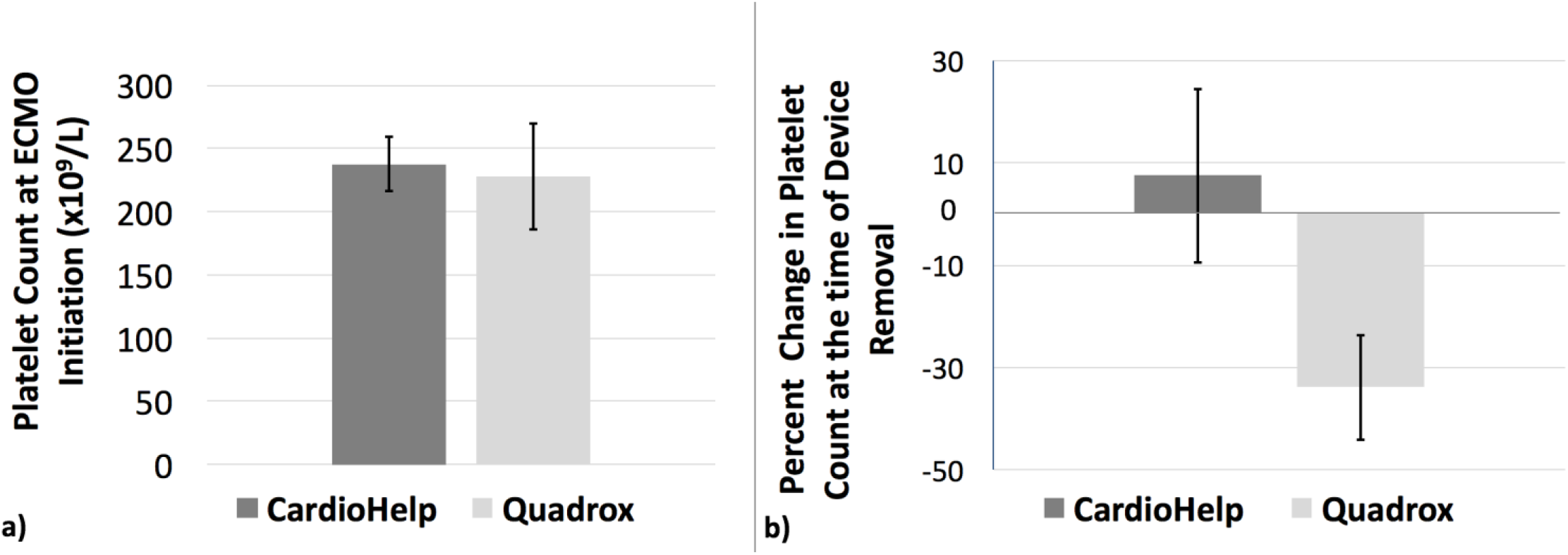
a) Average platelet count at time of ECMO initiation, b) Average percent change in platelet count at the time of ECMO discontinuation.

Resistance data are shown in **Figures 2a and 2b**. As seen in **Fig. 2a**, the Qx oxygenators have a more prominent early increase in resistance, whereas there is little change in resistance of the CH devices over the first 4 days. Around day 11, the average resistance of the Qx devices decreases, but this observation is due to 4 failed devices being removed, leaving behind only the most well-functioning devices and resulting in a selection bias. **Figure 2b** shows that when devices fail, they do so acutely with a sudden, exponential increase in resistance. Device survival data is depicted in **Figure 3**, which indicates that Qx devices meet the blood flow resistance failure criteria more quickly than CH oxygenators (p < 0.05).

**Figure 2:**
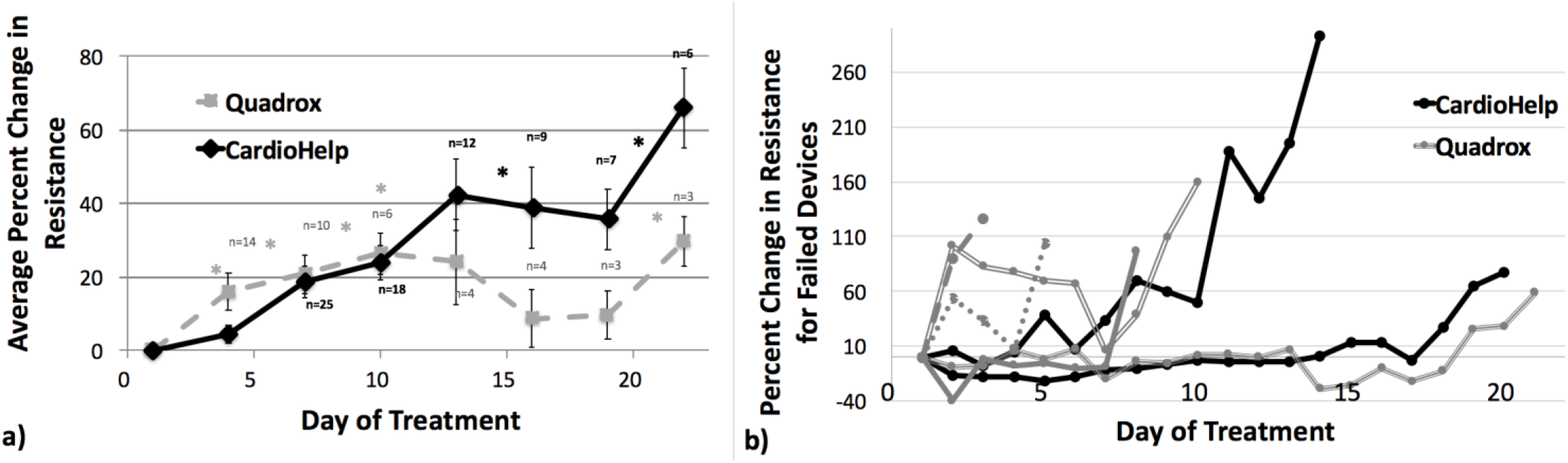
a) Percent change in resistance averaged over three days versus time. n = number of devices remaining. Asterisks (*) indicate device removal due to failure. b) Percent change in resistance over time for failed individual devices.

**Figure 3:**
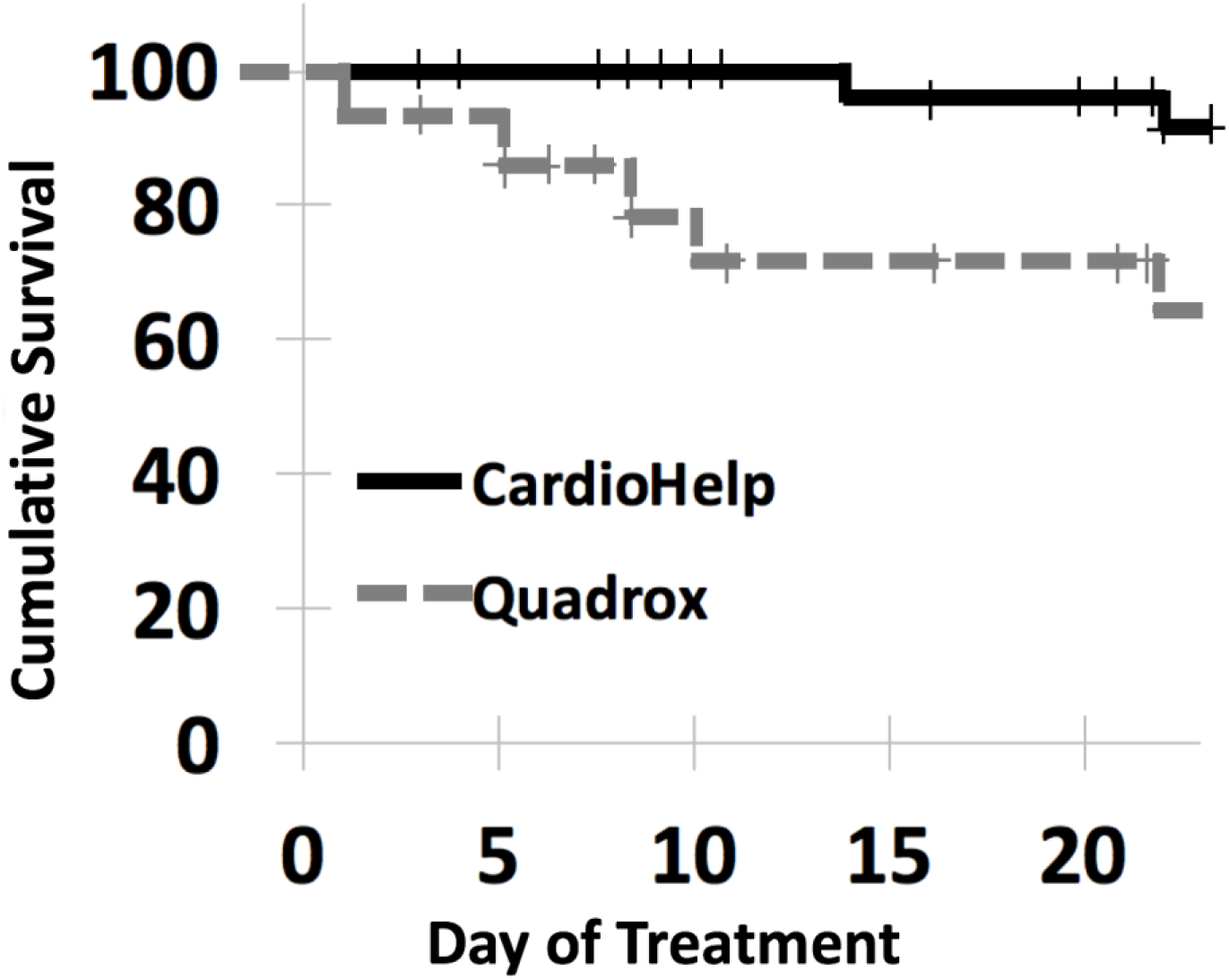
Device survival versus day on ECMO based on resistance failure criteria. Log-Rank, p = 0.014. Plus signs (+) indicate device removal due to weaning.

**Figure 4** shows the percent change in ΔC_O2_, with a greater decrease seen in the Qx group. These results parallel the resistance trends, with the CH maintaining performance for over 7 days, while the Qx devices experience a statistically significant decline in gas exchange by day 2 (p < 0.05) and a statistically significant increase in resistance by day 3 (p < 0.05).

**Figure 4:**
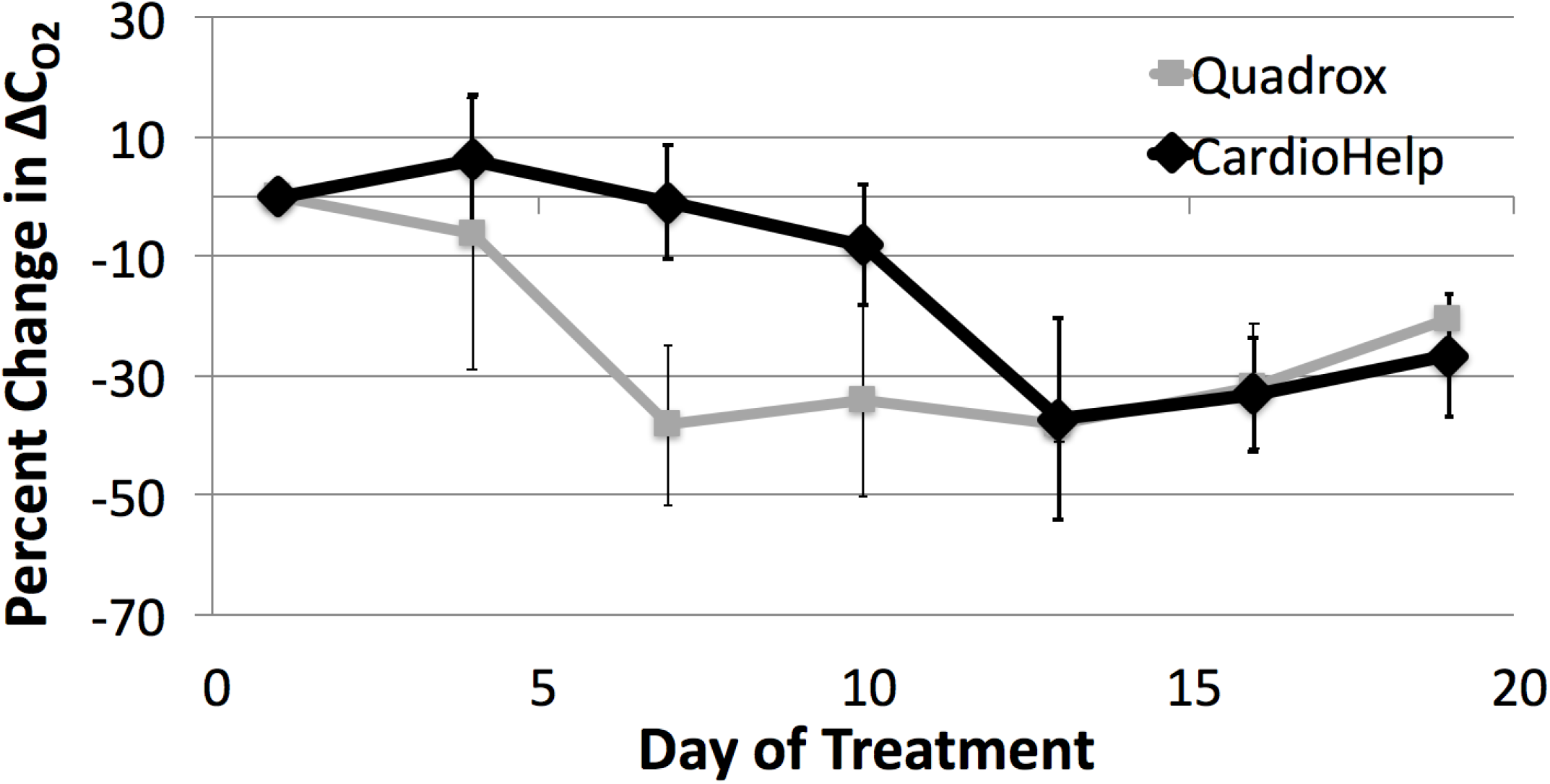
Percent change in inlet-outlet oxygen content difference (ΔC_O2_).

Probability densities of clot formation at the device inlets are seen in **Figure 5a**. As expected, clot formation is most significant at points most distal from the inlets. For the Qx group, there is increased clot at the inlet in the failed devices, coming predominantly from one corner, but also diffusely spread across the inlet face. In contrast, the two CH devices that failed had hardly any visible surface clot at the inlet. Progressive thrombus propagation over time is clearly evident when looking at averages of all devices removed over a specific range of days. With the CH devices, clot burden is strictly at the center of the device, farthest from all inlets, whereas in Qx devices, there is a more diffuse pattern of clot formation. **Figure 5b** shows probability densities of clot formation at the outlets. Unlike the inlet, clot formation at the CH outlet is indicative of failure, with failed devices showing a diffuse pattern of outlet clot formation. Similar to its inlet pattern, clot formation in the Qx outlet is more diffuse. Although surviving Qx devices had more clot formation at the corners, it likely didn’t affect performance significantly because those areas initially had reduced blood flow. Failed Qx devices, however, have a more diffuse clot pattern, indicating clot formation in the middle of the bundle. As expected, oxygenators associated with longer ECMO runs had increased clot burden at the device inlets. Unlike the Qx outlets which had increasing clot formation over time, the CH did not demonstrate substantial clot formation at its outlets.

**Figure 5:**
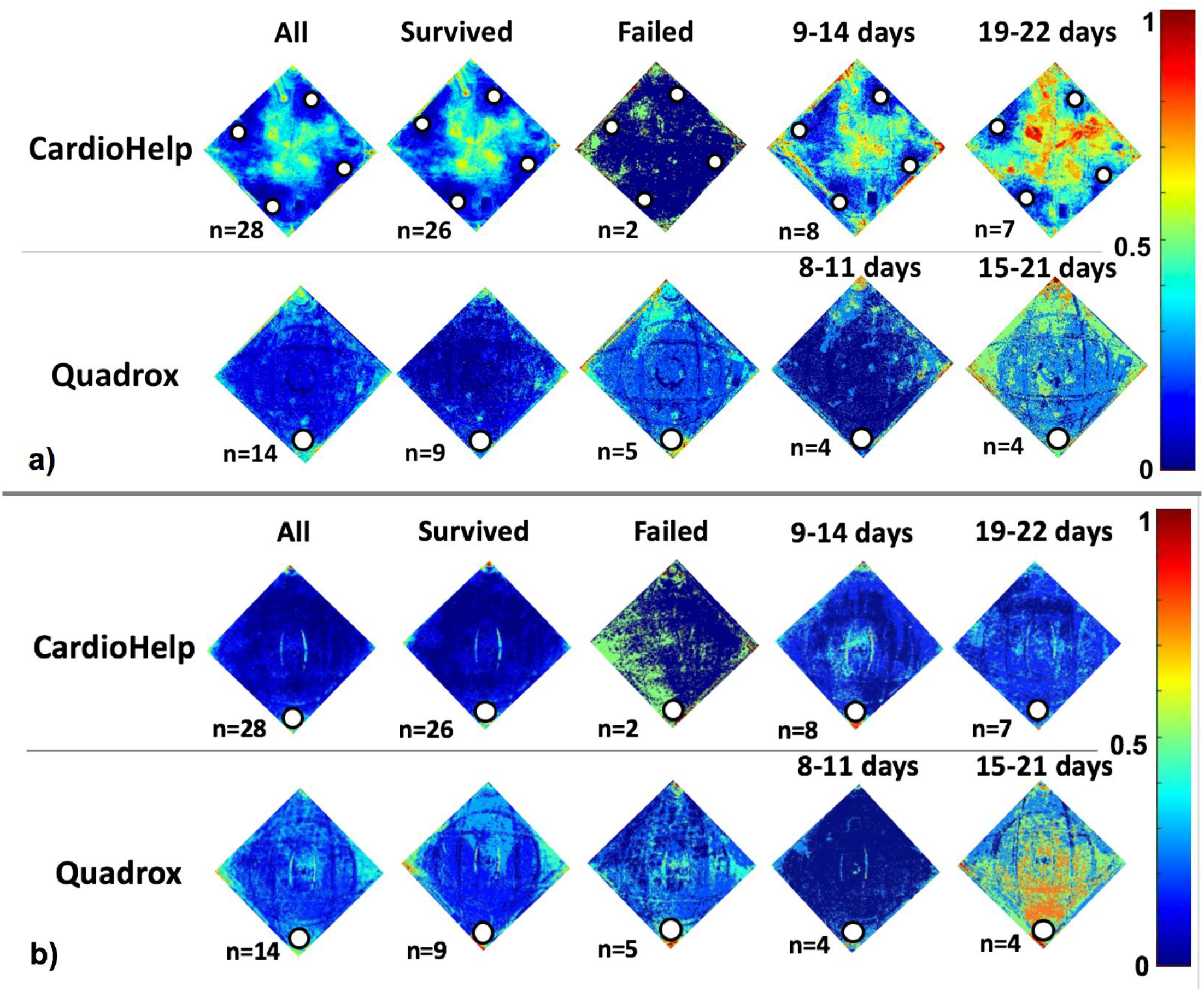
Probability density of clot formation. a) Device inlets. White circles indicate inlets. b) Device outlets. White circles indicate outlets. Color scale pictured: 0 (dark blue) indicates 0% clot frequency, 1 (dark red) indicates 100% clot frequency.

## Discussion

The development of an artificial lung as destination therapy is currently limited by clot formation. In order to prolong device lifetime in new designs, we must first understand patterns of clot formation in existing devices. To this end, we evaluated the time-course of thrombus development in two of the most commonly used clinical oxygenators.

Although the starting platelet counts aPTTs throughout the ECMO course, and circuit flow rates of the two patient populations were similar, there were distinct differences in various hematological and device characteristics over time. Overall, the Qx devices fail earlier and more frequently than CH devices, and when they fail, it is with a sudden, exponential increase in resistance. There was an early increase in resistance of the Qx oxygenators, with 4 out of 14 devices failing by day 11. Over the first 4 days, there was no change in resistance of the CH oxygenators, and it was not until day 15 that the first of two devices failed.

Platelet counts decreased significantly more in the Quadrox patients during the course of ECMO, likely due to deposition or destruction. Interestingly, this difference was seen even when sorting the patient populations into finer groups, such as indication for ECMO (**Table 2**). The only difference seen between the two groups that may be related is that the Qx patients required higher overall pump speed in revolutions per minute (RPM; CH 3020 ± 62, Qx 3413 ± 95, p = 0.013) to maintain similar average device flow between the two groups, (p = 0.69). All patients had a centrifugal pump. The Qx was paired with a Rotaflow or Centrimag in all cases, whereas the CH has an integrated centrifugal pump. The difference in RPMs between the CH and Qx devices remained regardless of pump type. The relationship between pump speed and pump shear stresses are likely to be different between the pumps, but this suggests that there may be more shear-induced platelet activation and loss in the Qx circuits than the CH circuits. Because these platelets travel directly from the pump to the oxygenator, this may contribute to the accelerated failure in the Qx oxygenators, however this would need be confirmed with further study.

As a measure of gas transfer performance, we calculated the inlet-outlet oxygen content difference (ΔC_O2_). This metric was chosen, rather than oxygen transfer, to eliminate some of the effect of device weaning since oxygen transfer is nearly linearly related with oxygenator blood flow rate. We identified an immediate decrease in ΔC_O2_ of the Qx oxygenators, which correlates with the increase in resistance seen in this group. Together, these suggest an increased early clot burden in the Qx oxygenators. On approximately day 13, the CH ΔC_O2_ declines substantially, which is around the same time the average resistance of this group exceeds that of the remaining Qx devices. After approximately 13 days, the remaining Qx devices and the CH devices show similar gas exchange function. Together, these results suggest that Qx devices are more prone to early failures in the first 10 days, but surviving devices function well thereafter relative to the CH devices.

The program used to analyze the images was able to clearly demonstrate clot formation patterns. In the CH devices, thrombus grows with time from the center of the device, farthest from the four inlets. It is likely, therefore, that the higher blood velocity at each of the four device inlets washes away clotting factors and thus maintains an open blood path, whereas the central region away from the inlets suffers from a mix of recirculation and stagnation, leading to rapid clot formation. Interestingly, the two CH devices that failed did not have substantial visible clot burden at the inlets, but had significantly more clot formation at the device outlet. It appears that the clot formation in the CH inlet does not interfere significantly with the normal blood flow path in this oxygenator, and thus little change in resistance is seen. However, CH devices with increasing resistance likely have more diffuse clot formation throughout the fiber bundle that grows from the inlet side toward the outlet with increasing concentrations of activated platelets and clotting factors. Notably, there were no differences found between the two patients with failed CH oxygenators, demographically or hematologically, that could explain why these devices failed more quickly.

In the Qx oxygenators, clot formation is more diffuse, although there is a general trend towards more clot formation at the point in the inlet manifold farthest from the blood inlet connector. This diffuse pattern of clot formation is likely the cause of the early failures in these devices. Moreover, unlike the CH oxygenators, the Qx outlets had a significant amount of diffuse clot that grew over time.

Overall, there were differences in patterns of clot formation, platelet consumption, resistance increases, and diminishing gas exchange efficiencies between the two oxygenators. The greater longevity of the CH oxygenators suggests that a more distributed, four-inlet approach is better than a single-inlet. Furthermore, although visual inspection is one of the considerations when deciding to switch out a device, our heat maps show that for the CH, only clot burden at the outlet correlates with device failure, which may be a subtle indicator given its understated visual manifestation. Additionally, percent increase in resistance appears to correlate with loss in gas exchange capabilities. Lastly, CH devices seem to provide more durable gas exchange than the Qx. Although not part of our study, the increased durability of CH may make it more cost effective than the Qx if a long ECMO run is anticipated due to less frequent need for oxygenator switch-outs. Although an economic analysis was not included in this study, it could form the basis for a break-even financial analysis that includes anticipated duration of ECMO support.

One limitation of this study is that our images only evaluate the inlets and outlets of the device and do not indicate the clot formation patterns on the interior. Future studies should consider performing similar assessment of the internal aspects of the fiber bundle, either by cutting devices open or using non-invasive imaging techniques. Other useful future studies to help explain the observed differences would include analysis of the relative rates of platelet activation by the pumps when isolated from their oxygenators and more detailed analysis of platelet activation and function and concentration of activated coagulation factors. One final limitation is that all information came from a single institution. Patient and circuit management differs based on institutional and clinician preference, which could influence the functional durability of the oxygenators.

## Data Availability

All data is available upon request.

## Acknowledgement

This work was supported by NIH grants 2R01HL089043 and R01HL089456 and The Richard Bartlett Foundation.

